# Validation of Automated Biventricular Myocardial Segmentation from Coronary Computed Tomographic Angiography for Multimodality Image Fusion

**DOI:** 10.1101/2021.03.08.21252480

**Authors:** Marina Piccinelli, Navdeep Dahiya, Russell D Folks, Anthony Yezzi, Ernest V Garcia

**Author notes:** **Corresponding Author** Marina Piccinelli, Ph.D., Emory University School of Medicine, Department of Radiology and Imaging Sciences, 1364 Clifton Rd, NE, Atlanta, Georgia, 30322.

## Abstract

**Purpose:** Image fusion strategies of myocardial perfusion imaging (MPI) and coronary CT angiography (CCTA) have shown increased diagnostic power. However, their clinical feasibility is hindered by the lack of efficient algorithms for the extraction of cardiac anatomy from CCTA datasets. The aim of this work was to validate our previously published algorithm for automated cardiac segmentation of CCTAs in a larger cohort of subjects while testing its application in clinical settings.

**Methods:** Three borders were automatically and manually extracted on sixty-three clinical CCTAs: left and right endocardia (LV, RV) and the biventricular epicardium (EPI). Impact of image resolutions and inter-operator variability on accuracy and robustness of automated processing were evaluated. Automated algorithm accuracy was assessed with the Dice Similarity Coefficient (DSC) and the surface-to-surface distance metric. Relevant quantities were compared for automated versus manual segmentations: LV and RV volumes, myocardial mass and LV myocardial mass.

**Results:** Lower resolution images offered an acceptable trade-off for accuracy and processing time (45 sec). DSC for LV, RV, EPI borders were 0.88, 0.80 and 0.89. Automated versus manual correlation coefficients for LV and RV vol, myo and LV mass were 0.96, 0.73, 0.84 and 0.67 with inter-operator agreement > 0.93 for three variables. Consistent and improved results were evidenced at higher resolutions.

**Conclusion:** Our algorithms allowed efficient automated cardiac segmentation from CT imagery with minimal user intervention, clinically acceptable times and accuracy. The reported results show promise for its use in a clinical environment, specifically in the context of image fusion.

## 1. Introduction

For decades the use of multimodality image fusion for the detection and assessment of Coronary Artery Disease (CAD) has been heralded as a powerful diagnostic strategy^1,2^. It allows the incorporation of anatomical and physiological information obtained from two or more imaging modalities and permits a more comprehensive portrayal of disease severity^3-6^. Even from a clinical perspective, it is not uncommon for a patient to be acquired with multiple modalities when the first test has produced ambiguous results. Yet, multimodality image fusion frameworks that can be used seamlessly in a clinical environment are not widely available, likely due to the lack of well-validated, robust and fast software tools for the automated quantification of anatomical acquisitions such as Coronary Computed Tomographic Angiography (CCTA).

Currently CCTA/nuclear myocardial perfusion imaging (MPI) fusion frameworks exist as research applications^6,7^ or in systems that do not rely on the explicit CCTA segmentation but rather on the visualization of the two datasets with volume rendering techniques^8^. In contrast, our team has been concentrating on the creation of frameworks that explicitly obtain a 3D representation of the heart anatomy^9-11^ to be displayed in combination with coronary trees and with functional quantitative assessments such as ischemic areas^12^, PET-derived absolute blood flow quantification^7^, and myocardium at risk^6^. Clinical implementation of our quantitative fusion technique requires that our software have access to accurately and quickly segmented nuclear perfusion and CCTA cardiac imagery. Extraction of the required nuclear information is readily available from our widely disseminated nuclear cardiology software (Emory approach^13^). Although many techniques have been reported for the whole heart segmentation from CCTA studies^14^, what has been lacking is a software package for the extraction of the required anatomic information from CCTA that is readily accessible to quantitative fusion algorithms.

We recently published our novel and improved methodology^15^ for the automated segmentation of a biventricular myocardium from clinical CCTAs that included positive preliminary results from a small population. However, crucial aspects for the evaluation of any segmentation algorithm that is being proposed for clinical use are degree of automation, speed, reliability, applicability in a medical environment and robustness. The aim of this work was to validate our proposed CCTA cardiac automated segmentation methodology in a larger cohort of subjects using more clinically significant validation parameters to better assess its reliability, accuracy, stability and speed while also testing the feasibility of its application in a clinical setting.

## 2. Materials and Methods

### 2.1 Study Population and Imaging Data

Sixty-three consecutive imaging datasets were selected from a database of clinical CCTAs available in our Nuclear Cardiology Laboratory at Emory University. The database was created with images acquired at different clinical and research centers as a result of previous collaborations^6^. The data was acquired between 2005 and 2016 with devices from different manufacturers (Siemens Somaton Definition, Siemens Sensation Cardiac 64, GE LightSpeed VCT and Philips Brilliance 64). The image acquisition was originally performed after review and approval by Emory University Institutional Review Board (USA), the Institutional Review Board of the University Hospital Val D’Hebron (Spain) and the Institutional Review Board of the Rambam Medical Hospital (Israel). Written informed consent was obtained from each patient in accordance with clinical guidelines on human research^6^. The current work retrospectively analyzes the previously collected images under a novel review and approval of the Emory University Institutional Review Board.

ECG-gated contrast-enhanced CT images were acquired following standard clinical guidelines after the injection of 60 mL of nonionic contrast agent at a rate of 4 mL/sec and saved in DICOM format. The best diastolic phase was selected for successive processing.

### 2.2 Volumetric Dataset Preparation and Manual Segmentation

To facilitate manual segmentation of cardiac CCTA images, datasets were re-oriented along the cardiac short-axis (SA) and resampled to create an isotropic volumetric dataset. A manual procedure common to nuclear cardiac studies processing was used to re-orient the CCTA transaxial images. Once re-oriented, the images were resampled with an isotropic image spacing equal to the original in-plane image resolution creating a volume of 512×512×512 voxels, indicated herein as High Res. To accelerate image processing and reduce computer memory requirements a scaled-down version of each image was created with a spacing 4 times the original one and a dimension of 128×128×128 voxels, indicated as Low Res. An in-house developed software was used for the manual segmentation performed by members of the team expert in cardiac anatomy. Three borders were identified on the volumes: the left (LV) and right ventricle (RV) endocardia and the biventricular epicardium (EPI). Manual segmentations were performed only on the higher resolution datasets. Binary images were constructed from the profiles and three-dimensional (3D) surface models extracted with the Marching Cube algorithm^16^. In Figure 1 the steps from transaxial images to 3D biventricular model are shown. The manual segmentations represented the ground truth for validating the automated results and the SA images were the only input to the segmentation code.

**Figure 1.**
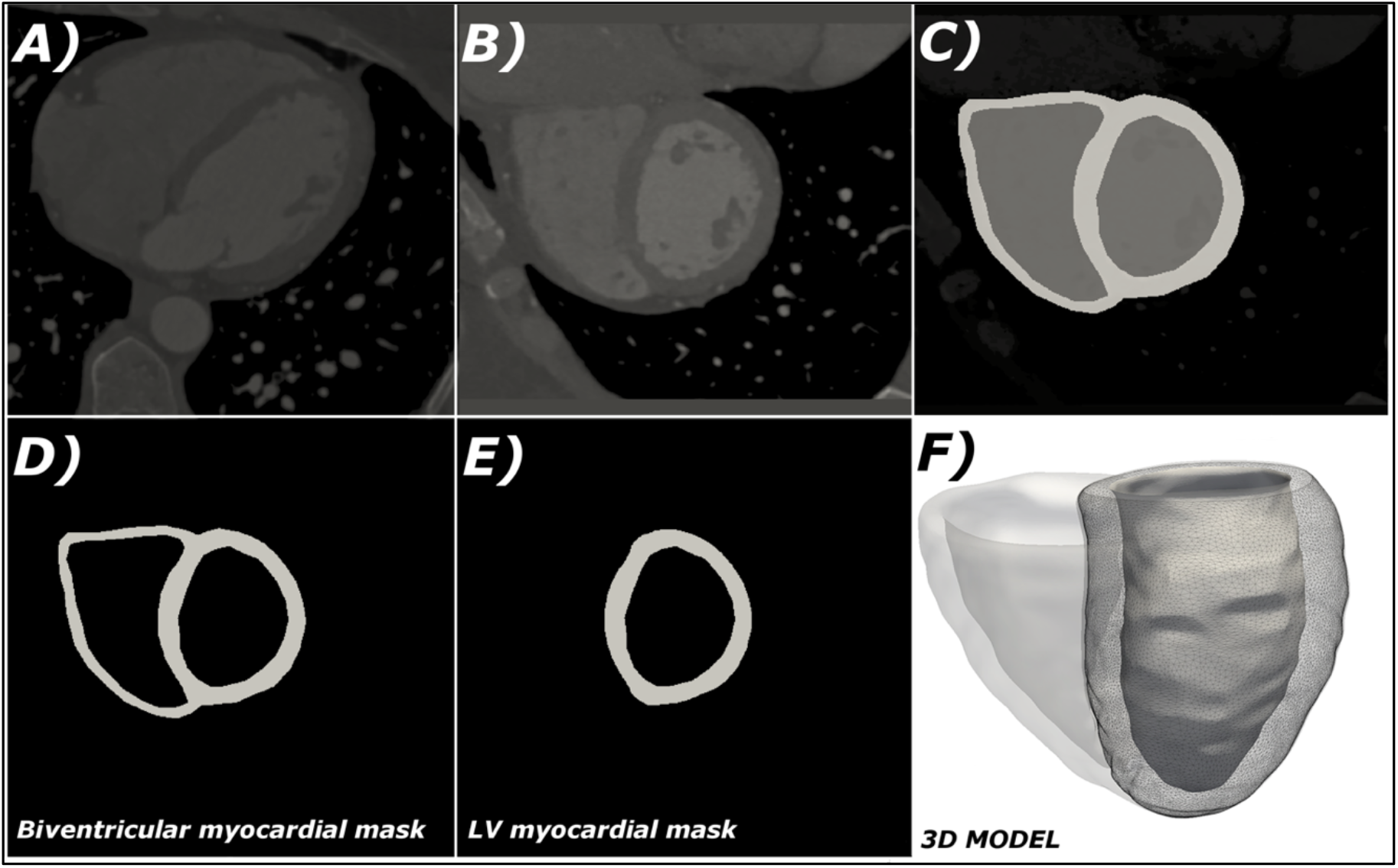
Reorientation of CCTA images and manual segmentation output: A) original transaxial orientation; B) re-orientation along the cardiac short-axis; C) manual segmentation mask identifying LV, RV chambers and myocardial tissue; D) biventricular myocardial binary mask; E) extraction of the LV myocardium; F) 3D model.

### 2.3 Automated Segmentation Algorithms

Our algorithms for the automated segmentation of the biventricular model of the heart from CCTA images were recently published^15^. While implementation details are available elsewhere, here we briefly describe the main features of the developed methodologies.

The algorithms can be considered a modification of the Chan-Vese techniques^17^ that represent an image as a piece-wise constant function where the evolution of the boundaries separating the different objects to be segmented depends on the statistics of the pixel intensities within each 3D region. Our proposed algorithms were customized to the problem of myocardial segmentation by introducing several high-level constraints, employing a binary (rather than constant) intensity model for the nearby background (BG) region, and making use of shape priors for the anatomical regions (LV, RV, EPI) that were obtained from an initial training phase. All these modeling ingredients are combined into a single integrated optimization problem iteratively solved within which the three boundaries are evolved simultaneously while the modeling constraints couple their evolution behavior (i.e. severely penalize overlap between boundary surfaces). Since in our formulation the borders do not evolve independently, weighting factors (indicated with w_BG_, w_LV_, w_RV_ and w_EPI_) could be assigned to the curve evolution to bias if needed the expansion/contraction of one region at the expense of another and to address cases of low contrast particularly between myocardium and RV. The weighting factors could theoretically be modified case by case, but in an effort to test the robustness and clinical applicability of the whole procedure, optimal setting values were identified in previous investigations (w_BG_ = 1.0, w_LV_ = 0.5, w_RV_ = 0.5 and w_EPI_ = 1.5) and used for all the analyzed cases as the default configuration for the algorithms. In the training phase, 10 randomly selected, manually segmented datasets were used to extract mean shapes and principal modes of shape variations for LV, RV and EPI. These training datasets guided the prospective segmentation of all other cases. A mean shape was computed for each anatomical structure from the training datasets and a principal component analysis (PCA) technique employed to extract shape priors which represented the individual variations in shape with respect to the mean.

### 2.4 Segmentation Software Utilization

The algorithms were incorporated into a stand-alone application written in C++ with a graphic user interface in Tcl/Tk that allows the operator to supervise and interact with the different modules of the procedure. In terms of interactions of the user with the code, once the weighting factors were fixed, the segmentation procedure was essentially fully automated. Only two inputs were required from the operator to model the background in the images (i.e. all the structures outside the myocardium): a representative intensity level for the bone structures and an average gray value for all other tissues. A display window enabled the user to pick points, visualize their intensity values and enter the two selected thresholds to be used in the running code.

Two sets of images were analyzed to investigate image resolution impact on automated segmentation: the 53 prospectively segmented sets at Low Res and a cohort of 20 High Res images. To assess inter-operator variability the Low Res images were processed by two users (indicated as U1, U2). The High Res images were processed only by the primary operator U1. The number of iterations was set to 100 for all cases in addition to the default weighting factors setting. Only the two background thresholds were selected case-by-case by the operator. The processing times were recorded.

The same ten datasets were used for the training phase at both high and low resolution to be properly employed in the prospective segmentations of the two cohorts but not included in the validation results.

### 2.5 Comparisons of Manual versus Automated Segmentations

A number of comparisons were performed to compare manually segmented images to the ones automatically obtained.

#### 2.5.1 Dice Similarity Coefficient

As manually created binary images of the LV, RV chambers and the biventricular EPI were considered the reference standard, the Dice Similarity Coefficient (DSC)^18^ was employed as one of the metrics to assess the algorithms accuracy. The DSC is an index of spatial overlapping commonly used to validate segmentation algorithms with binary images. It is computed as twice the number of pixels that belong to the segmented structure in both automated and manual masks over the sum of the number of pixels defining the structure in each of the mask. The DSC ranges between 0 and 1 (0 indicating no overlap and 1 complete overlap) and was computed for both the Low Res and the High Res cohorts. The manual binary masks were resampled to the same image resolution before comparisons in the Low Res group.

#### 2.5.2 LV and RV Volumes and Myocardial Masses

The following physiologically relevant quantities were computed for automated and manual segmentations: LV and RV chambers volume (Vol), and biventricular myocardial mass (Myo Mass) by subtracting the LV and RV binary images from the EPI multiplied by a density factor of 1.05 g/mL. Given the importance of the LV myocardium in cardiovascular diseases and MPI studies, an automated procedure for the removal of the right ventricle was used to obtain just the LV mass^7^ (LV Myo Mass) (see Figure 1 D-E).

3D surface models for the LV, RV and EPI were obtained and surface-to-surface distances computed between each couple of manual-automated anatomical structures. A surface-to-surface distance between triangulated meshes was defined as the Euclidean distance (in mm) of each point on the source surface mesh (automated) to the closest point in the target surface mesh (manual) along the direction of the local normal to the source surface. The average of surface-to-surface distance map for all points was computed for LV, RV and EPI surfaces. Figure 2 shows maps of the surface-to-surface distance calculation displayed for one of the analyzed cases.

**Figure 2.**
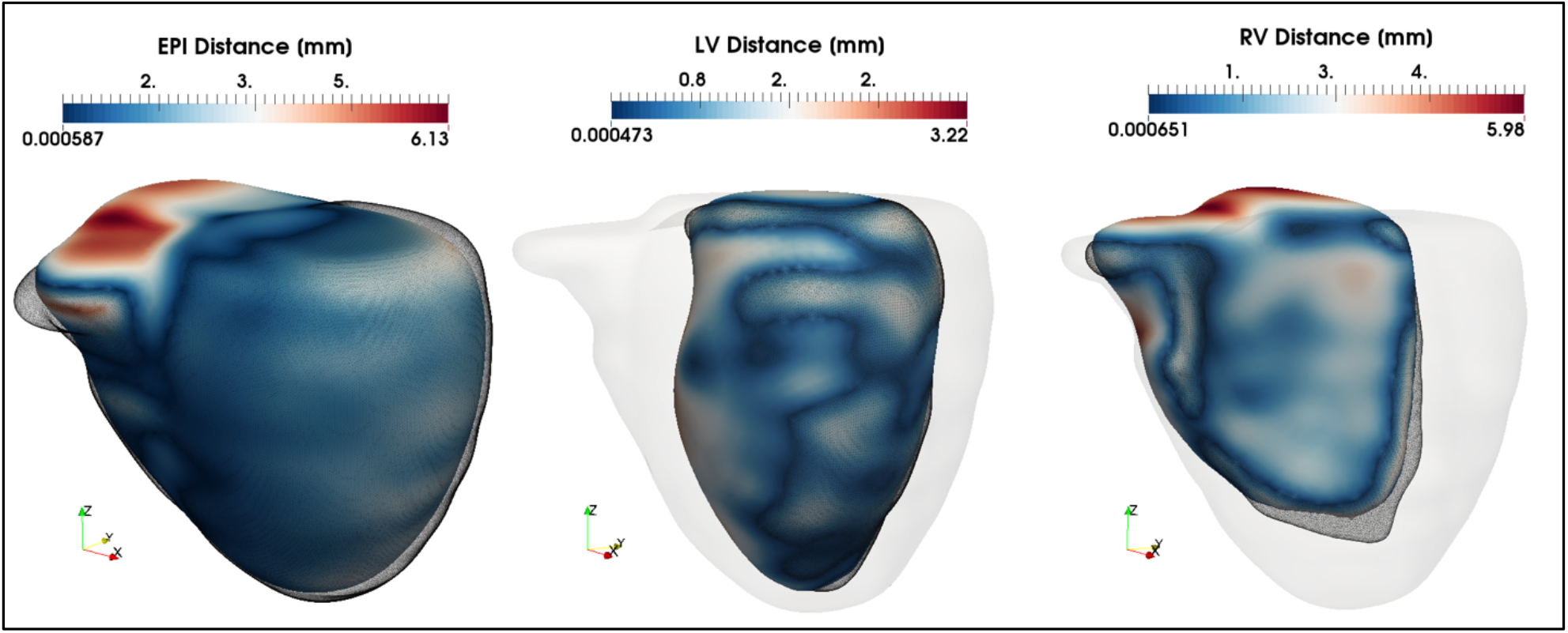
3D maps of the surface-to-surfaces distance between manually segmented borders (mesh in opaque) and automated ones (color-coded). The absolute distance between each couple of structures is used to color to models. For anatomical reference the manual epicardial model is also displayed in opaque light gray with the LV and RV chambers.

Volumes, masses and surface-to-surface distances were computed for all the analyzed datasets, for both operators and both image resolutions when available.

### 2.6 Statistical Analysis

Continuous data were expressed as mean ± SD and statistical significance was measured with Student t-test with a level of significance of p<0.05. The DSC was provided as mean ± SD. Pearson correlation coefficients (R) and the standard error of the estimate (SEE) were used to assess the performance of the automated segmentations in extracting LV and RV Vol, Myo and LV Myo Mass and to assess U1-U2 inter-operator variability. Percentage of relative errors of the automated measurements were calculated with respect to the manual ones. Surface-to-surface distances were provided as mean ± SD over the entire triangulated surfaces. Student t test were used to assess differences between operators processing and image resolutions with a level of significance of p<0.05.

## 3. Results

### 3.1 Image Preparation and Processing

Original in-plane image resolutions of the 63 cases used varied from 0.31 to 0.61 mm (mean 0.43 ± 0.06); the best diastolic phase ranged between 55% and 89% of the cardiac cycle.

### 3.2 Image processing time and robustness

The manual segmentation of all three borders required from 2 to 3 hours. The time required for image re-orientation and creation was consistently below 1-2 minutes. Of the 53 prospective studies, 44 visually converged on all three cardiac anatomic features using the weighting factors established during training while the remaining 9 required the operator to change the factors for the algorithm to converge. Thus, the assessment on the algorithms’ performance was based on the 44 studies where no interaction was required.

Automated segmentation was performed using a quad core 3.2 GHz Intel core i7 CPU with a 16 GB RAM computer (although the algorithms were not yet multithreaded and consequently unable to exploit the parallel processing capabilities). For the down sampled cases (Low Res) automated segmentation took an average of 0.45 secs per iteration favorably converging at 100 iterations (45 secs total). For the full resolution cases (High Res) automated segmentation took an average of 21.9 secs per iteration for a total of 36.5 min processing time.

### 3.3 Manual vs Automated Segmentations

The DSC was computed for all cases in the two image resolution cohorts. Mean values were as follows: 0.88 ± 0.04, 0.80 ± 0.08, 0.89 ± 0.04 respectively for the LV and RV chambers and biventricular EPI at Low Res (n=44); 0.91 ± 0.02, 0.85 ± 0.03, 0.89 ± 0.02 at High Res (n=20).

In Table 1 automated segmentations are compared to manual ones in terms of average values of LV and RV chamber volumes, and Myo and LV Myo Mass for the entire cohort of studies processed at Low Res (n=44) by primary operator U1. Figure 3 graphically depicts the performance of the automated segmentations for the four analyzed quantities. Mean percentage relative errors were 28, 31, 34 and 35% for LV and RV Vol, Myo and LV Myo Mass respectively and are shown in Figure 4 (left). While paired t tests exhibited statistically significant differences between manual and automated values for all variables, Pearson correlation coefficients showed excellent results for the LV endocardium detection (R = 0.96, SEE = 13.1) and progressively decreasing levels of accuracy for LV Myo Mass (R = 0.84, SEE = 21.7), RV Vol (R = 0.73, SEE = 27.8) and Myo Mass (R = 0.67, SEE = 29.3).

**Table 1.** Comparison of manual versus automated segmentations for the Low Res cohort. Images were processed by two users (U1, U2) with default algorithmic settings (n=44). LV Vol, left ventricle volume; RV Vol, right ventricle volume; Myo Mass, biventricular myocardial mass; LV Myo Mass, left ventricle myocardial mass; SD, standard deviation; SEE, standard error of the estimate; R, Pearson’s correlation coefficient.

| **n = 44** | **Manual** | **Automated (U1)** |  | **Automated (U2)** |  | **Inter-Operator Correlation** |
| --- | --- | --- | --- | --- | --- | --- |
|  | *mean ± SD* | *mean ± SD* | *R (SEE)* | *mean ± SD* | *R (SEE)* |  |
| **LV Vol**<br>[mL] | 96.9 ± 37.7 | 120.9 ± 43.9 | 0.96 (13.1) | 119.6 ± 43.6 | 0.95 (14.3) | 0.98 |
| **RV Vol**<br>[mL] | 105.4 ± 27.2 | 134.2 ± 40.3 | 0.73 (27.8) | 128.3 ± 38.4 | 0.67 (28.8) | 0.88 |
| **Myo Mass**<br>[g] | 170.3 ± 48.9 | 110.0 ± 39.0 | 0.67 (29.3) | 108.6 ± 38.3 | 0.72 (26.8) | 0.96 |
| **LV Myo Mass** [g] | 135.0 ± 43.8 | 85.5 ± 30.9 | 0.84 (21.7) | 81.7 ± 31.0 | 0.84 (16.8) | 0.93 |

**Figure 3.**
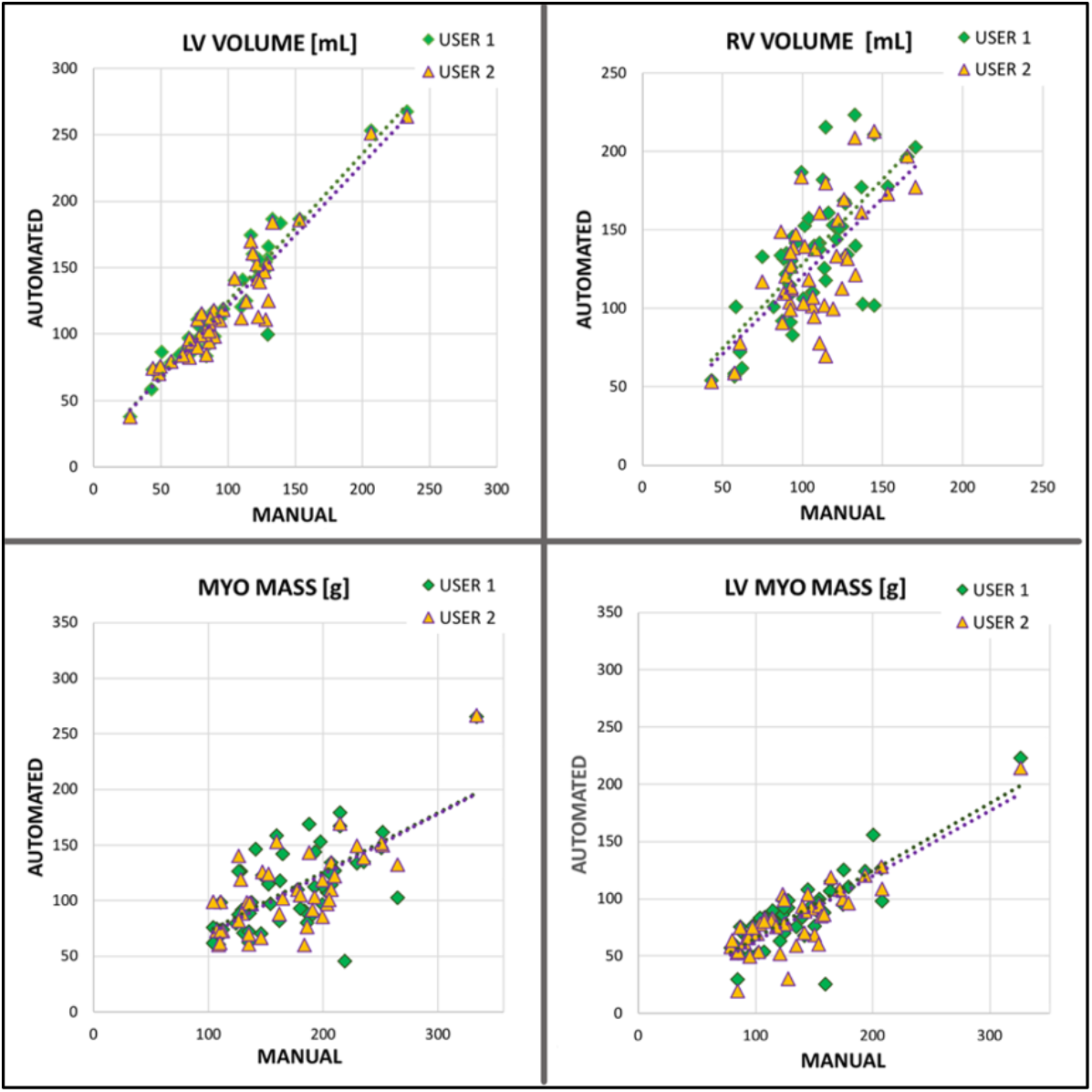
Performance of the automated algorithms in the extraction of LV and RV Volume, Myo Mass and LV Myo Mass compared to manual segmentations in the Low Res cohort (n=44). Two operators (User 1 and User 2) independently processed the data. Correlation coefficients reported in Table 1.

**Figure 4.**
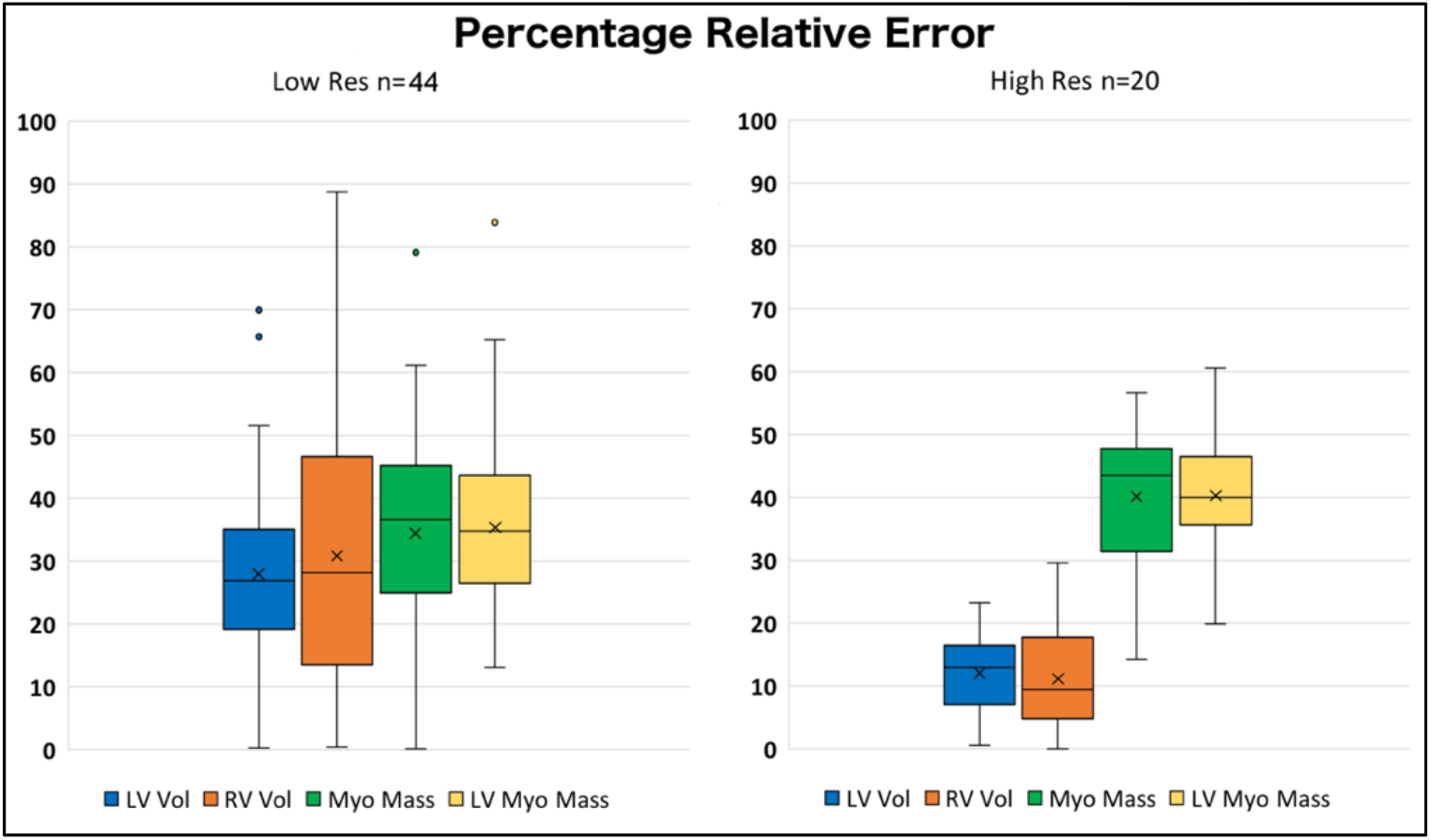
Distribution of percentage relative errors in the extraction of LV and RV Volume, Myo Mass and LV Myo Mass compared to manual segmentations for the Low and High Res cohorts. quantifications (p<0.05) but not in the computations of mass.

To assess inter-operator variability due to differences in selecting the two fiducial points required (background and bone), the Low Res cohort was processed by a second user U2. Table 1 reports mean values and correlation coefficients for U2 processing and inter-operator correlations for all measured variables. Statistically significant differences between U1 and U2 were found in the LV and RV volume quantifications (p<0.05) but not in the computations of mass.

Analogously to U1, the most accurate automated quantifications with respect to manual were LV Vol and LV Myo Mass with R = 0.95 and 0.84 respectively and decreasing performance for Myo Mass and RV Vol (R = 0.72 and 0.67). The inter-operator consistency was excellent with R ≥ 0.93 for three of the four measured variables.

In Table 2 and Figure 5 the manual vs automated comparisons are reported for the cohort of datasets processed at both high and low resolutions by U1 (n=20). The trend to improved accuracy is shown in all measurements for both R and SEE when processing at a higher image resolution. The averaged percentage relative errors for the High Res cohort were 12, 11, 40 and 40% for respectively LV and RV Vol, Myo and LV Myo Mass. Figure 4 (right) displays the error distribution over the entire cohort.

**Table 2.** Comparison of manual versus automated segmentations for the High Res and Low Res cohorts processed by primary user U1 (n=20). LV Vol, left ventricle volume; RV Vol, right ventricle volume; Myo Mass, biventricular myocardial mass; LV Myo Mass, left ventricle myocardial mass; SD, standard deviation; SEE, standard error of the estimate; R, Pearson’s correlation coefficient.

| **n = 20** | **Manual**<br>*mean ± SD* | **Automated (U1)** |  |  |  |
| --- | --- | --- | --- | --- | --- |
|  |  | **High Res**<br>*mean ± SD* | **R (SEE)** | **Low Res**<br>*mean ± SD* | **R (SEE)** |
| **LV Vol**<br>[mL] | 120.1 ± 40.9 | 134.6 ± 48.6 | 0.99 (7.4) | 139.4 ± 49.7 | 0.96 (14.1) |
| **RV Vol**<br>[mL] | 120.6 ± 24.0 | 119.3 ± 23.6 | 0.75 (16.0) | 149.4 ± 34.5 | 0.59 (28.6) |
| **Myo Mass** [g] | 199.8 ± 33.0 | 120.0 ± 33.6 | 0.68 (25.5) | 107.0 ± 31.8 | 0.52 (28.0) |
| **LV Myo Mass** [g] | 148.4 ± 27.7 | 88.0 ± 18.3 | 0.71 (13.3) | 82.2 ± 23.1 | 0.63 (18.4) |

**Figure 5.**
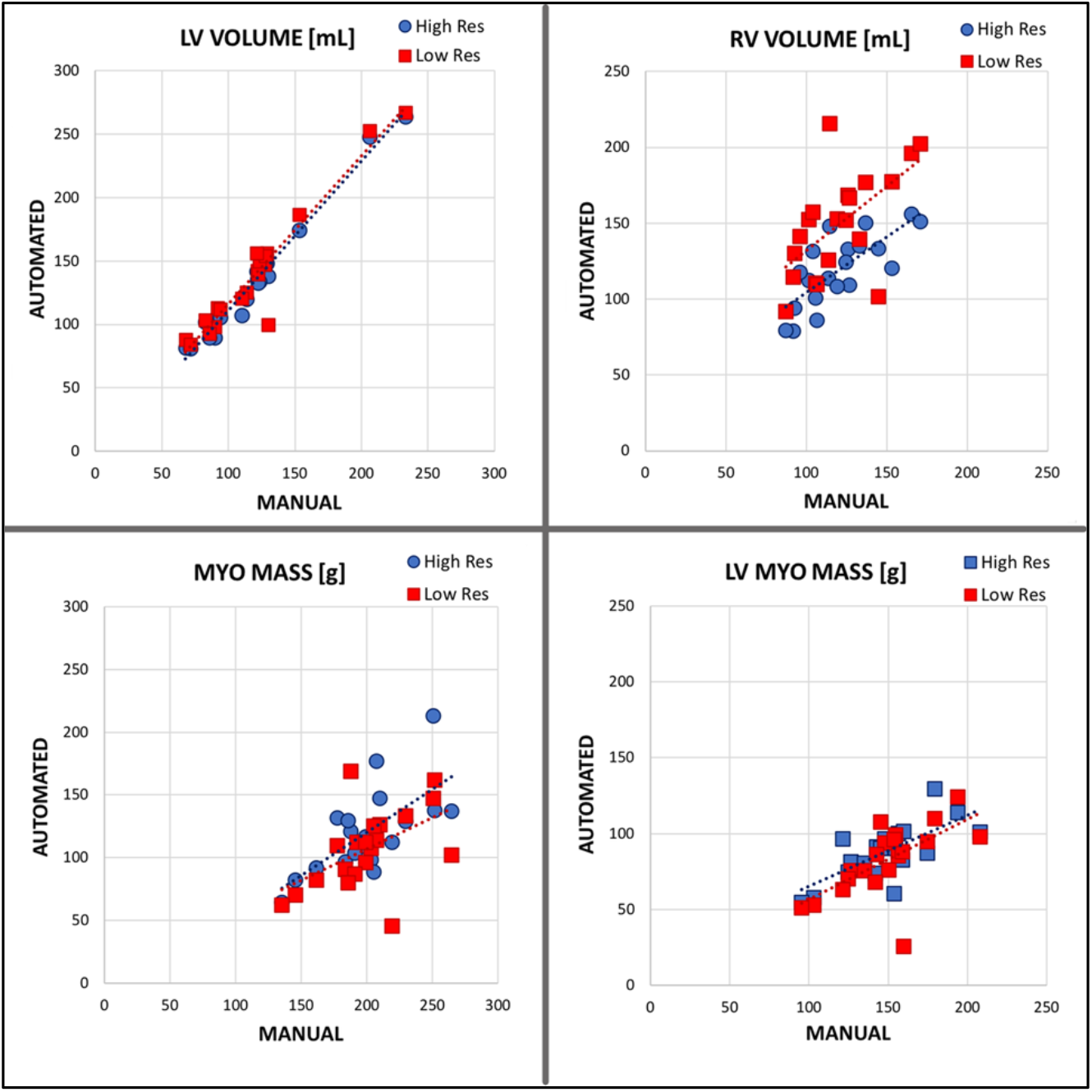
Performance of the automated algorithms in the extraction of LV and RV Volume, Myo Mass and LV Myo Mass compared to manual segmentations at both high and low image resolution (n=20).Correlation coefficients reported in Table 2.

Table 3 reports the analysis of the 3D surface-to-surface distance between each couple of automated-manual borders, LV and RV endocardia and the biventricular EPI. Results of U1 processing confirmed the highest accuracy of the LV chamber identification with a 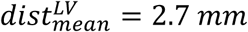 and slightly higher values for EPI and RV borders (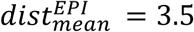 and 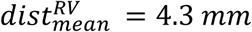). These assessments were supported by results from U2 processing: analogous values were found for the LV and EPI surfaces (p-values>0.05), while differences were shown in the identification of the RV chamber (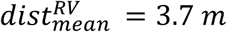; p-value<0.05) by the second operator. The maximum distance values could greatly increase on a case by case basis (in fact as much as 4.5 cm), but these values were consistently located at the base of the heart or even further out with respect to where the manual segmentations stopped. In Figure 6 two examples are displayed of automated vs manual 3D models. In Figure 6-B the automated models notably expanded outside of the manual segmentation domains. The usage of images with higher resolution improved the results and decreased the automated versus manual mean distances for all 3D surfaces (Table 3, column III and IV). The segmentations of LV and RV chambers were significantly more accurate (p-value<0.01), while only a trend toward lower errors could be evidenced for EPI borders. The significant decrease in maximum distance values also substantiated the better performance of the algorithms in case of high resolution images.

**Table 3.** Surface-to-surface distances calculated for the Low Res cohort (n=44) processed by two users (U1, U2) and in the subgroup of datasets processed at both image resolutions (n=20) by primary user U1. LV Endo, left ventricle endocardium; RV Endo, right ventricle endocardium; Heart Epi, biventricular epicardium. Values reported as mean ± SD [min -max] range; **‡** p-value < 0.05; *, ** p-value < 0.01

| Surface-to-Surface Distances |  |  |  |  |
| --- | --- | --- | --- | --- |
|  | Low Res (n=44) |  | U1 Processing (n=20) |  |
|  | U1 | U2 | High Res | Low Res |
| **LV Endo**<br>[mm] | 2.7±1.0 [3.8-24.6] | 2.6±1.1 [3.6-21.0] | 1.7±0.4 [3.8-11.9] * | 2.6±1.0 [3.8-22.7] * |
| **RV Endo**<br>[mm] | 4.3±2.4 [5.9-45.0] ‡ | 3.7±1.6 [6.1-27.8] ‡ | 2.3±0.5 [5.5-14.2] ** | 3.9±1.5 [5.9-30.2] ** |
| **Heart Epi**<br>[mm] | 3.5±1.58 [7.3-37.8] | 3.2±1.4 [6.6-31.8] | 3.3±0.5 [8.0-17.0] | 3.8±1.5 [7.3-30.0] |

**Figure 6.**
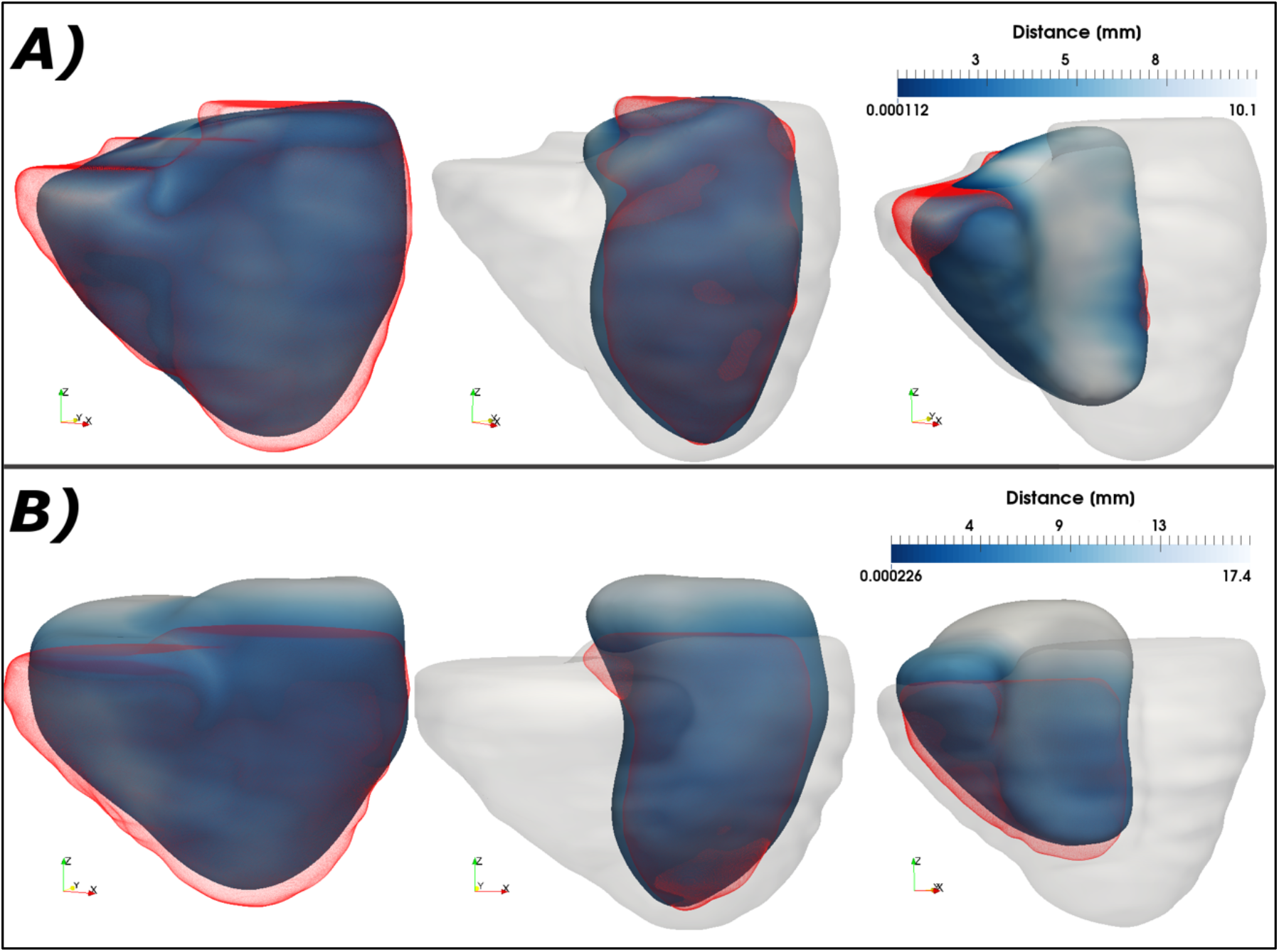
Display of two of the analyzed cases comparing automated and manual 3D models for the biventricular epicardial surface (left), LV (mid) and RV (right) chambers. The automated models are color-coded with the absolute surface-to-surface distance, and the manual ones are shown in opaque red. For anatomical reference the manual epicardial model is also displayed in opaque gray with the LV and RV chamber.

## 4. Discussion

In this study, we determined the favorable accuracy of the automated detection of biventricular myocardial surface from CCTA acquisitions in 44 clinical patients. The accuracy was determined by comparing the automated surfaces to those hand-traced by experts. These comparisons were done in terms of: a) physiologically important cardiac anatomic parameters (LV and RV volumes, myo and LV myo mass) determined by the automated technique, b) the boundary distances measured between automatic and hand-traced surfaces and c) the DSC for the LV and RV chambers and biventricular EPI.

Our results show that by down-sampling the high-resolution CT images we could extract the required automated segmented anatomic features at a clinically acceptable processing time (45 secs) with a reasonable trade-off for accuracy. Our best results were found in the segmentation for the LV chamber and biventricular epicardium and our least accurate the RV chamber. The high inter-operator correlation coefficients also substantiated the robustness and reliability of the methodology even when used by less experienced users.

In the course of this testing, overall favorable performance resulted in the segmentation of the left and right ventricular boundaries away from the apex and base structures, but a lesser performance was noted near the apex as well as in underestimated septum thickness between the ventricles as evidenced by the errors on the LV myo mass assessment (Fig 4). Analysis and testing at Georgia Tech have linked much of this underperformance to uncertainties and inconsistencies in capturing the structures at the base, resulting in larger errors in the distances between the boundaries at these locations (Fig 6, B). Work is in progress that focuses on improving these shortcomings.

Many other automated methods exist for segmenting anatomic cardiac features from CCTA and have been reported in excellent reviews. Notably, a recent review [14] compares results from 10 methods applied to the same 60 CT studies acquired in a clinical environment with manual segmentation for comparison. The evaluated methods included deep-learning (DL) and multi-atlas (MAS) segmentations. Although difficult to compare due to differences in datasets and exact processing tasks, our results compare favorably with their average results for DSC (0.87 DL, 0.86 MAS vs. ours 0.88 (High Res), 0.86 (Low Res)), surface-to-surface distance (mm) (1.84 DL, 3.3 MAS vs. ours 2.4 (High Res), 3.5 (Low Res) and processing time (range: 0.22 sec – 21 min vs. our down-sampled average: 45 secs).

The present manuscript presents a crucial benchmark for the development and extension of our image fusion framework in order to provide radiologists and researchers with an easy-to-use application to analyze, quantify and visualize different imaging modalities into a single integrated display. While the results favorably compare with other techniques and methodologies, an additional validation will be necessary to assess whether the accuracy of the present automated segmentation is sufficient to guarantee the correct alignment of CCTA and cardiac MPI (either SPECT or PET) and consequently the fusion of anatomic structures, namely the patient-specific coronaries, to functional data for diagnosis purposes. A similar study was previously published by our team on our animal model experiments^19^.

While the relative errors between manual and automated segmentation may appear significant (Fig 4), it should be noted that their impact on our registration algorithms^20^ and consequently on image fusion has not been evaluated yet. As our multimodality registration algorithm starts with the alignment of the LV chambers and uses the additional features (the biventricular myocardial mass and the RV chamber) to correct for rotations around the LV short-axis, the influence of such errors may not be sizable given the much more modest image resolution of the nuclear images. With this work, the authors do not wish to claim that at the present stage of development the algorithms could be used to reliably extract all anatomical features from CCTA images, but that automated MPI/CCTA image fusion can finally become a *clinical reality*. Furthermore, notably CCTA protocols focus on the visualization of the LV structures, as the most important part of cardiac function, not necessarily on the RV and biventricular masses, which explains the extremely variable image quality evidenced for the right ventricle and the less reliable segmentation results.

### Limitations

A limitation of our work is that it used the interactively hand-traced CCTA biventricular meshes as a reference standard. Although these surfaces were defined by clinical experts, manual methods are subjective.

We also did not compare directly our automatic CT contours to those from other automated methods^14^. This was not done for a number of reasons. As discussed above, a large number of different segmentation solutions have indeed been proposed through the years to segment anatomic information from CT acquisitions. Our future plans are to compare our segmentation to those available to us using the same image datasets but, since no methodology has emerged as a definitive reference for comparisons’ sake and the long-term goal of this project is automatic nuclear MPI/CTA fusion, the direct availability of CCTA-derived surfaces is of paramount importance hence the need to develop our own integrated procedure.

Our automated algorithms have been developed to take as input data the CCTA best diastolic phase as the time point at which cardiac structures are less susceptible to motion artifacts. Although specific testing has not been performed on this issue, different cardiac phases could be theoretically used once the training step had been adjusted to the new cardiac time point.

## 5. Conclusions

Our proposed algorithms allowed efficient segmentation of LV, RV and EPI from CCTA clinical datasets with minimal user intervention in clinically acceptable times and accuracy. The reported results show promise for its use in a clinical environment and specifically in the context of multimodality image fusion.

## Data Availability

Patients imaging datasets were selected from a database of clinical CCTAs available in the Nuclear Cardiology Laboratory at Emory University. The database was created with images acquired at different clinical and research centers as a result of previous collaborations from 2005 and 2016.

## Conflict of Interest

Some of the authors (EVG, RDF) receive royalties from the sale of the Emory Cardiac Toolbox and have equity positions with Syntermed, Inc. The terms of these arrangements have been reviewed and approved by Emory University in accordance with its conflict of interest policies. The remaining authors did not report any conflicts of interest.

## Source of Funding

This work was supported by National Institutes of Health (NIH) Grant R01 HL143350 (PI: EV Garcia). The content is solely the responsibility of the authors and does not necessarily represent the official views of the NIH.

## Notes

### Author Declarations

The CCTA image acquisition was originally performed after review and approval by Emory University Institutional Review Board (USA), the Institutional Review Board of the University Hospital Val D Hebron (Spain) and the Institutional Review Board of the Rambam Medical Hospital (Israel). Written informed consent was obtained from each patient in accordance with clinical guidelines on human research. The current work retrospectively analyzes the previously collected images under a novel review and approval of the Emory University Institutional Review Board.

